# Genome sequencing reveals the impact of pseudoexons in rare genetic disease

**DOI:** 10.1101/2024.12.21.24318325

**Authors:** Georgia Pitsava, Megan Hawley, Light Auriga, Ivan de Dios, Arthur Ko, Sofia Marmolejos, Miguel Almalvez, Ingrid Chen, Kaylee Scozzaro, Jianhua Zhao, Rebekah Barrick, Nicholas Ah Mew, Vincent A. Fusaro, Jonathan LoTempio, Matthew Taylor, Luisa Mestroni, Sharon Graw, Dianna Milewicz, Dongchuan Guo, David R. Murdock, Kinga M. Bujakowska, UCI-GREGoR Consortium, Changrui Xiao, Emmanuèle C. Délot, Seth I. Berger, Eric Vilain

## Abstract

**Purpose:** Advancements in sequencing technologies have significantly improved clinical genetic testing, yet the diagnostic yield remains around 30-40%. Emerging sequencing technologies are now being deployed in the clinical setting to address the remaining diagnostic gap.

**Methods:** We tested whether short-read genome sequencing could increase diagnostic yield in individuals enrolled into the UCI-GREGoR research study, who had suspected Mendelian conditions and prior inconclusive clinical genetic testing. Two other collaborative research cohorts, focused on aortopathy and dilated cardiomyopathy, consisted of individuals who were undiagnosed but had not undergone harmonized prior testing.

**Results:** We sequenced 353 families (754 participants) and found a molecular diagnosis in 54 (15.3%) of them. Of these diagnoses, 55.5% were previously missed because the causative variants were in regions not interrogated by the original testing. In 5 cases, they were deep intronic variants, all of which led to abnormal splicing and pseudoexons, as directly shown by RNA sequencing. All 5 of these variants had inconclusive spliceAI scores. In 26% of newly diagnosed cases, the causal variant could have been detected by exome sequencing reanalysis.

**Conclusion:** Genome sequencing overcomes multiple limitations of clinical genetic testing, such as inability to call intronic variants and technical limitations. Our findings highlight pseudoexons as a common mechanism via which deep intronic variants cause Mendelian disease. However, they also reinforce that reanalysis of exome datasets can be a fruitful approach.

## Introduction

The widespread application of clinical genetic testing has tremendously increased the rate at which rare genetic diseases are being diagnosed and new gene-disease associations are identified. Despite this progress, the diagnostic yield of clinical genetic testing remains 30-40% [1, 2]. There are several reasons for this plateau. For many conditions, the causal genes are still unidentified. Even for conditions with known causal genes, interpreting the clinical significance of variants can be challenging. This is because there are a large number of variants with insufficient information pertinent to their pathogenicity, known as variants of uncertain significance (VUS). In addition, technical limitations of commonly offered clinical tests can result in missed variants, for example in regions that are not represented in exome sequencing such as deep intronic and untranslated regions. Epigenetic factors that affect gene expression but are not interrogated by standard testing potentially add to the complexity.

Molecular sequencing technologies not traditionally used in the clinical setting are now being deployed in order to overcome these challenges. These include the increasing use of genome sequencing (GS) as well as RNA sequencing. The latter can be especially useful for directly evaluating the functional consequences of intronic variants and variants in splice regions, since despite advances in computational predictors of splicing disruption (such as SpliceAI and Pangolin [3, 4]) it often remains difficult to determine if these variants are pathogenic or not.

The University of California, Irvine (UCI) - GREGoR site, part of the GREGoR (Genomics Research to Elucidate the Genetics of Rare Diseases) Consortium, has been evaluating the diagnostic utility of these newer approaches. Our study has been enrolling participants whose clinical presentation strongly suggests a genetic cause, but who remain without a molecular diagnosis, despite having undergone conventional genetic testing (gene panels, chromosomal microarray, exome sequencing - ES).

Here, we describe our center’s experience from short-read GS of a cohort of 353 such families (754 participants). To help disambiguate previously or newly detected variants (VUS or single heterozygous pathogenic variants in recessive disease genes), we deployed additional methods, including RNA sequencing. We also provide an overview of our cohort and focus on lessons learned from informative, successfully solved cases.

## Methods

### Participants

The Mendelian Genomics Research Center was launched in 2021, between Children’s National Hospital, Invitae, and more recently UCI. The study was approved by the Institutional Review Board (IRB) of Children’s National Hospital (Pro00015852). Study participants included probands with a suspected Mendelian condition and prior non-diagnostic clinical testing, such as gene panels, chromosomal microarray, or ES. Testing on additional family members was performed when available. Individuals were enrolled over a three-year period, from July 2021 to April 2024, through referrals by physicians and collaborators, or self-referral via our website (https://research.childrensnational.org/labs/pediatric-mendelian-genomics-research-center). Written informed consent was available in both English and Spanish and was obtained for all families by study staff or through a study-specific automated chatbot [5]. Workflows for remote enrollment and sample collection were established, eliminating the need for participants to travel. Following consent, buccal swabs and/or peripheral blood samples were collected from probands and their relevant available family members. Some participants provided clinically generated genome data for reanalysis [6]. In addition, our study population involved two collaborative cohorts, one focusing on aortopathy and another on dilated cardiomyopathy; for these their prior genetic testing may not have been as comprehensive or well documented. Phenotypic categories are described in **Table 1**.

### Short-read Genome Sequencing

Blood and buccal samples were sent to Invitae® laboratories where they were processed and underwent genome sequencing using Illumina NovaSeq 6000. Sequencing files were subsequently aligned to the hg38 human genome assembly with bwa-mem using soft clipping enabled for supplementary alignments (−Y), processed at 120,000,000 input bases per batch for reproducibility (−K) [7] and genotyped using Google DeepVariant [8]. Structural variant calling was performed with Illumina Manta [9]. Alignments and variant call files are available on the NHGRI Analysis Visualization and Informatics Lab-space (AnVIL; AnVIL Portal) on the GREGoR Consortium workspace.

### Short-read RNA sequencing

Blood samples were preserved in PAXgene tubes processed with the WatchMaker Genomics RNA library prep kit with the Polaris depletion module to create stranded and ribosomal/globin-depleted libraries. They then underwent RNA sequencing using the Illumina NovaSeq 6000 S4 flowcells generating 100-200 million 150-bp read pairs. Reads were aligned to hg38 using STAR (v2.7.10a) [10] with the GENCODEv41 annotation. We employed the basic two-pass mode and allowed up to 3 mismatches and a minimum aligned length of 100 bp. Alignments were visualized using integrative genomics viewer (IGV) [11-14]. Alignments are available on AnVIL (AnVIL Portal).

### Mini gene assays

A minigene splicing assay was performed using a mini-gene split Green Fluorescent Protein (GFP) construct [15], in which N- and C-terminal parts of the *GFP* gene were separated by *SMN1* (MIM# 600354) introns 7 and 8 (NM_000344.4). Reference and mutated gene fragments containing 1000 bp of *DBT* (MIM# 248610) intron 10 surrounding the NM_001918.5:c.1282-4218G>A variant flanked with 30 bp vector homology arms were synthetized (TWIST Bioscience, USA) and cloned into the mini-gene construct (Gibson Assembly Master Mix, New England Biolabs). After Sanger sequencing verification of all constructs, they were transfected into HEK293 cells (Lipofectamine 3000, Thermo Fisher Scientific). Forty-eight hours post transfection, total RNA was extracted from the transfected cells (RNAeasy Mini Kit, Qiagen) and cDNA was generated using random hexamer primers (SuperScript IV Synthesis Kit, Thermo Fisher Scientific). Subsequently, the minigene transcripts were amplified from the cDNA using primers specific to the split *GFP* fragments (F: 5’-CACACTGGTGACAACATTTACATAC-3’; R: 5’-GAAATCGTGCTGTTTCATGTGATC-3’). The PCR products were column-purified (DNA Clean & Concentrator-5, Zymo Research) and analyzed with next-generation amplicon sequencing (MiSeq, Illumina, OGI Genomics Core). The splicing pattern analysis was performed by aligning the sequence reads to the hybrid reference of the split *GFP* construct containing the *DBT* intron 10 (STAR Aligner) [16] and visualizing the reads in (IGV) [11-14]. The same process was followed to test the effect of the *HFE* (MIM# 613609) VUS found in participant PMGRC-124-124-0.

### Variant prioritization and analysis

Phenotypic data for probands was collected via a manual review of electronic medical records or provided by collaborating researchers. The phenotypes of each proband were converted to Human Phenotype Ontology (HPO) terms.

Variants were reviewed by expert curators using the MOON^**TM**^ software. MOON implements a natural language processing approach to prioritize variants in genes that are likely to be related to an individual’s phenotype based on published literature [17, 18]. Cases were reviewed with the goal of identifying pathogenic, likely pathogenic, and VUS in known disease genes consistent with the phenotype of the proband. In addition to sequence and structural variants prioritized by MOON, rare sequence variants in the following categories were reviewed: those expected to result in loss of function, *de novo* variants, compound heterozygous or homozygous variants, hemizygous variants, variants classified as likely pathogenic or pathogenic in ClinVar, and variants in genes associated with conditions with a high clinical overlap for the patient.

Cases were considered “solved” when a disease-causing variant or multiple disease-causing variants in a gene associated with a condition consistent with the phenotype of the proband were detected. The number and phase of variants needed to be consistent with the inheritance pattern of the condition. For example, a single likely pathogenic or pathogenic variant in an autosomal dominant condition or biallelic likely pathogenic or pathogenic variants in an autosomal recessive condition were needed for a case to be considered “solved”. Cases were considered “probably solved” when one or more of the variants were classified as a VUS but there was a strong phenotypic overlap with the clinical presentation of the patient. Cases were considered “partially solved” if likely pathogenic or pathogenic variants consistent with the inheritance pattern of the condition were detected, but the result only explained part of the proband’s phenotype.

Finally, we note that analysis is ongoing; the data reported in this manuscript are as of October 1, 2024.

## Results

### Genome Sequencing leads to a molecular diagnosis in 15.3% of previously unsolved cases

A total of 353 families underwent GS during the designated period; demographic information is shown in **Figures 1a and 1b**. Our study included several different family structures; 41.9% (n=148) were proband-only cases, 13.3% (n=47) were duos or duos plus another family member (duo+), 43.3% (n =153) were trios or trios with another family member (trio+) and 1.4% (n=5) consisted of other family combinations (**Figure 2a**). This accounts for a total of 754 genomes analyzed.

**Figure 1.** **(A), (B)** Sex and race distribution of probands that underwent genome sequencing.

**Figure 2.** **(A)** Family structures corresponding to the participants **(B)**. Trio/Trio+ had the highest diagnostic yield (21%) whereas Duo/Duo+ and Proband only families reached a diagnostic yield of 13% and 10% respectively (p = 0.03, chi-square test).

A genetic diagnosis was found in 54 (15.3%) cases (solved/probably solved; Supplemental Table 1). As expected, the diagnostic yield was higher in trios/trios+ compared to duos/duos + and proband-only families (21% vs 13% and 10%; **Figure 2b**). This difference was statistically significant (p = 0.03, chi-square test).

In addition, 6 cases (1.7%) were partially solved; that is, the identified variant could explain part, but not the entirety, of the individual’s phenotype. Furthermore, we found a candidate variant in approximately 9% of the cases that remained unsolved (27 out of the 299; Supplemental Table 2).

To better understand why the diagnosis was previously missed in the 54 solved/probably solved cases, we examined the type of prior testing these participants had undergone (**Figure 3a**). In 26% (14 out of 54) of solved cases, the causal variant had been identified by initial testing but was not reported either because the gene-phenotype association was unknown at the time of that testing or because the variant was not classified as pathogenic at the time (**Figure 3b**). Of note, out of the 353 probands in total, 143 had previously undergone ES (**Figure 3c**). Below, we focus on cases where initial testing did not interrogate the genomic region harboring the causative variant.

**Figure 3.** **(A)** Types of prior testing undergone by participants that were deemed solved or probably solved (*1 patient had methylation studies in addition to panel testing; ** 3 patients had mitochondrial testing in addition to ES/GS and panel testing). **(B)** Reasons diagnosis was previously missed on clinical genetic testing. Limited original testing includes gene panels that lacked the causative gene or did not detect the causative variant due to its location in a genomic region not covered by the test. New information/Reclassification refers to variants reclassified due to new papers, or inclusion of ClinGen updates to the ACMG guidelines such as increasing strength for high revel scores, or further phenotypic evidence which allowed further variant criteria to be applied. (**C**) Pie chart showing the percentage of probands that had undergone exome or genome sequencing; percentages are calculate across the entire cohort (not only solved cases).

### Diagnoses in uninterrogated regions of original testing highlight pseudoexons as a common mechanism by which deep intronic variants cause disease

In the majority of solved/probably solved cases (55.5%; 30 out of 54 cases), the diagnosis was missed before because the original testing method was unable to call variants in the region harboring the causative variant. For example, there were 13 cases where panel testing did not include the causal gene, and 13 cases where the causative variant was non-coding. In one case, a synonymous coding-region variant was predicted by spliceAI to lead to intron retention in *HFE*. This intron retention event leading to a stop-gain was confirmed using a mini gene assay (Supplemental Figure 1).

With respect to non-coding causative variants, there were 5 cases with variants affecting deep intronic regions. In all of these, the causative variant altered the canonical splicing pattern via the inclusion of a pseudoexon (**Table 2**). Variant pathogenicity was revealed by RNAseq, which directly demonstrated the effect at the transcript level. For example, one case was due to an intronic variant in *PEX1*(MIM# 602136), in a patient with a known biochemical diagnosis of Zellweger Spectrum Disorder and a heterozygous known pathogenic variant NM_000466.3:c.2916del p.(Gly973fs) in *PEX1*. The pseudoexon was revealed by RNAseq (Supplemental Figure 2). Another example was a newborn female who was biochemically diagnosed with thiamine-responsive maple syrup urine disease (MSUD) based on the same diagnosis in her sister. However, all prior genetic testing (panel testing of 9 genes, *ASL* (MIM# 608310) single-gene testing, and ES) was negative. GS revealed a homozygous deep (−4218) intronic single nucleotide variant predicted to lead to aberrant splicing of *DBT*, a gene known to cause MSUD type II (MIM# 620699), which can sometimes be thiamine-responsive. RNAseq again confirmed the presence of a pseudoexon, establishing the molecular diagnosis (**Figure 4a**). This variant was also confirmed by minigene assay (**Figure 4b-e**). In another case we found a deep intronic variant in a case of Coffin-Siris, which we validated by showing that the episignature profile of this individual as determined using CpG methylation status from long-read sequencing, matches that of individuals with Coffin-Siris syndrome, as described in detail separately [19]. The case with a deep intronic variant in *HADHB* (MIM# 143450) is also separately described [20]

**Figure 4.** **(A)** RNA sequencing reads visualized with sashimi plot to indicate splicing events showing the pseudoexon inclusion in *DBT* in the proband (PMGRC-658-658-0) and the affected sibling as a result of a deep (−4218) homozygous intronic single nucleotide variant. **(B-E) Mini-gene splicing assay for the *DBT* c.1282-4218G>A variant. B)** Reference and c.1282-4218G>A variant *DBT* intron 10 fragments were cloned into a mini-gene split *GFP* construct, in which N and C-terminal parts of the *GFP* gene were separated by *SMN1* introns 7 and 8 (NM_000344.4). The construct was expressed in HEK293 cells for 48 hours followed by mRNA extraction, cDNA generation and NGS amplicon sequencing. Black arrows on the construct image indicate primer placement. **C)** RT-PCR gel electrophoresis demonstrates inclusion of the pseudoexon in both the wild type and reference transcript. **D)** Sashimi plots of reference and variant hybrid DBT-GFP constructs demonstrating a preferential pseudoexon (10B) inclusion in the sequences containing the c.1282-4218G>A variant. **E)** Inclusion of pseudoexon 10B leads to a premature stop codon at the end of the exon, which most likely escapes nonsense-mediated decay and leads to a truncated protein with a C-terminus different by 16-residues.

We found that, in all five cases with a pseudoexon, both the acceptor gain and the donor gain spliceAI scores fell within a range generally considered inconclusive with regards to impact on splicing (range 0.16 - 0.43 for acceptor gain scores; 0.19 - 0.55 for donor gain scores). This observation, while preliminary, suggests that the combination of such acceptor gain and donor gain scores may be a marker for variants leading to this splicing alteration. Of note, commonly used pre-calculated spliceAI scores limit their search to 50 bp from the variant, and therefore in each of these examples the new splice acceptor would not have been detected with pre-calculated spliceAI scores alone. It required a direct calculation of a spliceAI score with a search of at least 500 bp from the variant. Pangolin scores were generally higher than spliceAI scores.

Four cases harbored variants in the spliceosomal non-coding RNA *RNU4-2* (MIM# 620823) and contributed to the discovery of ReNU syndrome (MIM**#**620851). As described in detail in Chen *et al*. [21] variants in this gene represent a new, surprisingly common etiology of neurodevelopmental disorders, explaining approximately 0.4% of such cases. All four variants we identified fell within the 18bp region, which was shown to be enriched for pathogenic variants, of which three are the most common recurrent indel [20].

Finally, an informative example in which the causal variant was missed due to variant filtering parameters driven by limitations of ES was a woman in her 30s with glomuvenous malformations, conductive hearing loss, and midline malformations. Previous testing included a cancer 38-gene panel, *GLA* single-gene testing, ES, and mitochondrial testing; all were negative. GS revealed a known pathogenic exonic frameshift variant NM_053274.3:c.157_161del p.(Glu52_Lys53insTer) in exon 3 of *GLMN* (MIM# 601749; Glomuvenous Malformation Syndrome; MIM#138000). Upon review of the previous clinical ES, we discovered that this same variant was detectable in the original data. However, the variant had not been reported as the exon was excluded from the lab’s clinically validated regions due to a propensity for sequencing artifacts in this exon likely due to regional sequence complexity or homology.

Clinicians ordering clinical exome sequencing may not be aware of the coverage limitations of clinically relevant genes or exons important for the testing indication.

### Detection of structural variants

We also identified structural variants not detected by previous testing. This includes a complex rearrangement of *OCA2* (MIM# 611409) with deep intronic breakpoints [6] and deletion of the second exon of *CREBBP* (MIM# 600140) in a child with features of Rubinstein-Taybi syndrome 1 (MIM**#**180849) and previous negative ES. It is important to note that while ES has made strides in detection of structural variants, copy-neutral variants and single-exon deletions may still not be detected.

### Syndromic phenotypes had the highest solve rate

When examining the solve rate separately for each phenotypic category, the largest number of diagnoses were obtained in individuals with syndromic phenotypes (32 solved cases out of 128 total cases; 25%), while the solve rate in non-syndromic cases was 9.8% (**Figure 5A**). When further stratifying non-syndromic cases according to the affected organ system (**Figure 5B**), individuals whose phenotype was categorized as ‘Cardiovascular’ were the most likely to receive a diagnosis in our entire cohort (7 out of 19, 36.8 %). All but one of these individuals who received a diagnosis were affected by dilated cardiomyopathy and were part of a legacy cohort. The genes we found were not known to be associated with dilated cardiomyopathy at the time of testing, which explains why they were initially missed.

**Figure 5.** (**A**) The solve rate for syndromic versus non-syndromic cases. (**B**) The solve rate stratified by phenotypic category for the non-syndromic cases.

## Discussion

Our study provides a characterization of the potential of short-read GS, combined with other confirmatory analyses such as RNAseq, to yield genetic diagnoses for previously undiagnosed cases. GS provides a molecular diagnostic advantage over ES in that it can detect variants in regions not interrogated by ES such as introns or non-coding genes, while RNAseq can assist in establishing the pathogenicity of intronic splice-altering variants by directly demonstrating the impact on the RNA product. Here, using a combination of GS and RNAseq we discovered pseudoexons as a common (among intronic variants) molecular mechanism with a loss-of-function impact via perturbed splicing.

Regarding causative variants in non-protein coding genes, while some examples of disease-causing non-coding RNAs were previously known, this number is now expanding thanks to the implementation of GS. Recently discovered examples include the aforementioned *RNU4-2*, encoding the U4 small nuclear RNA (snRNA) component of the major spliceosome [21, 22], and the long non-coding RNA *CHASERR* (MIM# 620993), which was shown to cause a novel syndromic neurodevelopmental disorder [23].

Going forward, we envision that improvements in our ability to evaluate the impact of variants in regions traditionally missed by conventional genetic testing will continue to increase the diagnostic yield of GS. It is important to recognize, however, that in some cases it may be cost-effective to reanalyze ES data instead of resorting to GS, especially given advances in variant interpretation driven by better predictive models of variant effects and an expanded understanding of tolerated genetic variation. Indeed, many causal variants in our study could have been detected by a reanalysis of ES data, using new information provided by variant reclassification or improved analytic pipelines; a finding that is in alignment with results from previous studies [24-26]. In addition, testing of other family members can also aid in variant reinterpretation by showing if a variant is *de novo* or if it segregates with the phenotype in the family. Finally, we speculate that newer long-read sequencing technologies will help further close the diagnostic gap by capturing variants not captured by short-read technologies such as certain structural variants and repeat expansions [27].

## Data Availability

Data is available in the GREGoR workspace in AnVIL (AnVIL Portal).

https://anvilproject.org/

## Data Availability

Data available in the GREGoR workspace in AnVIL (AnVIL Portal).

## Acknowledgements

We thank the participants and referring physicians for participating in this study.

## Ethics Declaration

This study was approved by the Children’s National Hospital Institutional Review Board (IRB) under protocol Pro00015852. Informed consent was obtained from all participants as required by the IRB.

## Author Contributions

Conceptualization: G.P., S.I.B, C.X., E.C.D., E.V.; Data curation: G.P., M.H., I.C., K.S., J.Z.; Formal analysis: G.P., M.H., I.C., K.S., J.Z., I.d.D., L.A. K.M.B. A.K.; Resources: M.A., S.M., S.M., N.A.M, M.T., L.M., S.G., D.,M., D.G., D.R.M.; Software: S.I.B., L.A.; Writing original draft: G.P.; Writing-review & editing: G.P., S.I.B, C.X., E.C.D., E.V., M.H., J.L., R.B.

