## Supplementary material for "Genome sequencing reveals the impact of pseudoexons in rare genetic disease": Tables

Table 1. Definitions of Phenotypic Categories and number of cases for each category.

| Multisystem Syndromic Disorders  (#128) | Proband presenting with symptoms affecting multiple organ systems, often accompanied by distinctive facial features. |
| --- | --- |
| Non-syndromic Disorders  (#225) | Proband with conditions that predominantly affect a single organ system, without significant involvement of other systems. |
| Subcategories | |
| Neurodevelopmental Disorders  (#66) | Probands exhibiting conditions that primarily affect the development and function of the nervous system, including the brain, spinal cord, and peripheral nerves. This category encompasses disorders impacting cognitive function, behavior, motor skills, and neurological processes. |
| Cardiovascular Disorders  (#19) | Proband with primary involvement of the heart and blood vessels, such as structural heart defects, cardiomyopathies, arrhythmias, and vascular diseases. |
| Connective Tissue & Skeletal Disorders  (#98) | Proband with symptoms affecting bones, joints, and connective tissues such as skin, tendons, and ligaments. |
| Other  (#42) | Proband who do not clearly fit any of the aforementioned categories. |

Table 2. Participants with pseudoexon inclusion.

| Participant ID | Gene  (MIM ID; transcript ID) | Variant (c.) | Variant (g.) | SpliceAI score | | Distance to variant | | Pangolin score (Gain) | ACMG Classification (ACMG Evidence Criteria) |
| --- | --- | --- | --- | --- | --- | --- | --- | --- | --- |
|  |  |  |  | Acceptor | Donor | New Acceptor | New Donor |  |  |
| PMGRC-146-146-0 | *ARID1B* (#614556; NM_001374828.1) | c.3235+700C>G | [NC_000006.12:g.157167885C>G](https://www.ncbi.nlm.nih.gov/nuccore/NC_000006.12?report=graph&search=NC_000006.12%3Ag.157167885C%3EG) | 0.31 | 0.19 | -143 bp | -5 bp | 0.54 | Pathogenic (PS4, PM2, PS2, PP4, PVS1) |
| PMGRC-220-220-0 | *HADHB* (#143450; NM_000183.3) | c.1390-515_1390-499del | NC_000002.12:g.26289403-26289469del | 0.27 | 0.27 | -85 bp | 27 bp | 0.35 | LP (PVS1, PM2, PM3) |
| PMGRC-332-332-0 | *CRPPA* (#614631; NM_001101426.4) | c.789+973C>G | NC_000007.14:g.16307550G>C | 0.43 | 0.55 | 118 bp | 5 bp | 0.46 | LP (PM3, PM2, PVS1) |
| PMGRC-403-403-0 | *PEX1* (#602136; NM_000466.3) | c.1359+601A>G | NC_000007.14:g. 92513247T>C | 0.30 | 0.54 | 168 bp | 0 bp | 0.36 | LP (PVS1, PM3, PM2) |
| PMGRC-658-658-0 | *DBT*  (#248610; NM_001918.5) | c.1282-4218G>A | NC_000001.11:g. 100200640C>T | 0.16 | 0.20 | 161 bp | 3 bp | 0.66 | LP (PM2, PVS1) |

SpliceAI scores were calculated using the spliceAI lookup tool with a maximum distance of 500 bp. *LP* likely pathogenic
