## Supplemental Tables for "Genome sequencing reveals the impact of pseudoexons in rare genetic disease"

Supplemental Table 1. Solved/Probable solved cases

| **Participant ID** | **Phenotypic Category** | **Gene**  (MIM ID; transcript ID) | **Variant (c. or n.)** | **Variant (p.)** | **Variant (g.)** | **Inheritance** | **Patient Phenotype** | **Reason diagnosis was missed on previous testing** | **Associated Disorder (OMIM)** | **ACMG Classification** | **ACMG Criteria used** |
| --- | --- | --- | --- | --- | --- | --- | --- | --- | --- | --- | --- |
| PMGRC-13-13-0 | Syndromic | *PTHLH* (#168470; NM_198965.2) | c.4C>T | p.(Gln2*) | NC_000012.12:g.27969491G>A | Unknown | Pseudohypoparathyroidism, Chiari 1 malformation, shortened metacarpals | Limited Original Testing | Brachydactyly, type E2 (#613382) | Pathogenic | PM2, PVS1, PP4 |
| PMGRC-32-32-0 | Other | *FLG* (#135940; NM_002016.2) | c.1297_1298del  c.2282_2285del | p.(Asp433fs)  p.(Ser761fs) | NC_000001.11:g.152313589_152313590del  NC_000001.11:g.152312601ACTG | Paternally inherited,  Maternally inherited | Eczematoid dermatitis, numerous arcuate plaques with erythematous border, perivascular and interstitial dermatitis with neutrophils on biopsy, ichthyosiform xerosis | Limited Original Testing | Ichthyosis vulgaris (#146700) | Pathogenic  Pathogenic | PS4, PM3, PVS1, PM2  PM3, PVS1, PM2, PP5 |
| PMGRC-43-43-0 | Syndromic | *HECTD1* (#618649; NM_015382.4) | c.2082C>G | p.(Phe694Leu) | NC_000014.9:g.31150072G>C | De novo | Hypotonia, oral dysphagia, macrocephaly, autism spectrum disorder, somatic overgrowth syndrome, hypothalamic obesity, tube feeding, esotropia, stereotypic movements, abnormal brain MRI, dysmorphic facial features | New Information/Reclassification/Candidate gene now linked to syndrome | HECTD1- neurodevelopmental disorder (under review) | LP | PM2, PS2, PP2 |
| PMGRC-46-46-0 | Syndromic | *PPP1R3F* (#301104; NM_033215.5) | c.554_555del | p.(His185fs) | NC_000023.11:g.49270423_49270424del | Maternally inherited | Hypotonia, tube feeding, autism spectrum disorder, speech/language delays, developmental delay | New Information/Reclassification/Candidate gene now linked to syndrome | X-linked PPP1R3F neurodevelopmental disorder [27] | Pathogenic | PM2, PVS1, PP4 |
| PMGRC-88-88-0 | Syndromic | *NARS2* (#612803; NM_024678.6) | c.749G>A  2 exon deletion | p.(Arg250Gln)  p.? | [NC_000011.10:g.78493136C>T](https://www.ncbi.nlm.nih.gov/nuccore/NC_000011.10?report=graph&search=NC_000011.10%3Ag.78493136C%3ET)  NC_000011.10:g.78476711_78486357del | Paternally inherited  Maternally inherited | Critically ill neonate with multisystemic disease. A few months later, new findings of interstitial lung disease and refractory seizures | Limited Original Testing | Combined oxidative phosphorylation deficiency 24 (**#**616239) | LP  Pathogenic | PS4, PM2, PM3  1A, 3A, 2E, 4L |
| PMGRC-101-101-0 | Syndromic | *CBL* (#165360; NM_005188.4) | c.1259G>A | p.(Arg420Gln) | [NC_000011.10:g.119278541G>A](https://www.ncbi.nlm.nih.gov/nuccore/NC_000011.10?report=graph&search=NC_000011.10%3Ag.119278541G%3EA) | Nonmaternal | Hemihypertrophy, nystagmus, club foot | Previously known | Noonan syndrome-like disorder with or without juvenile myelomonocytic leukemia (**#**613563) | Pathogenic | PS4, PS3, PM2, PM5, PP3 |
| PMGRC-114-114-0 | Connective tissue & Musculoskeletal disorders | *ACAN* (#155760; NM_001369268.1) | c.4753del  c.757+4A>G | p.(Asp1585fs)  p.? | NC_000015.10:g.88857338del  NC_000015.10:g.88841871A>G | Paternally inherited,  Maternally inherited | Severe scoliosis, spondyloepimetaphyseal dysplasia, genu valgum, short stature | Previously known | Spondyloepimetaphyseal dysplasia, aggrecan type (**#**612813) | Pathogenic  VUS | PP4, PVS1, PM2, PM3  PM2, PP3, PM3 |
| PMGRC-124-124-0 | Syndromic | *HFE* (#613609; NM_000410.4) | c.187C>G,  c.892G>A | p.(His63Asp)  p.(Glu298Lys) | [NC_000006.12:g.26090951C>G](https://www.ncbi.nlm.nih.gov/nuccore/NC_000006.12?report=graph&search=NC_000006.12%3Ag.26090951C%3EG)  NC_000006.12:g.26092960G>A | Assumed biparental | Liver disease, hyperammonemia, encephalopathy, hyperbilirubinemia | Limited Original Testing | Hemochromatosis, type 1 (**#**235200) | Pathogenic  VUS | PS3, PP5  PM3, PM2, PP3 |
| PMGRC-146-146-0 | Syndromic | *ARID1B* (#614556; NM_001374828.1) | c.3235+700C>G | p.? | [NC_000006.12:g.157167885C>G](https://www.ncbi.nlm.nih.gov/nuccore/NC_000006.12?report=graph&search=NC_000006.12%3Ag.157167885C%3EG) | De novo | Developmental delay, gross motor delay, abnormal brain MRI, mixed receptive-expressive language disorder, dysmorphic facial features | Limited Original Testing | Coffin-Siris syndrome 1 (**#**135900) | Pathogenic | PS4, PM2, PS2, PP4, PVS1 |
| PMGRC-148-148-0 | Syndromic | *RNU4-2* (#620823; NC_000012.12) | n.64_65insT | NA | [NC_000012.12:g.120291839_120291840insA](https://www.ncbi.nlm.nih.gov/nuccore/NC_000012.12?report=graph&search=NC_000012.12%3Ag.120291839_120291840insA) | De novo | Hypotonia, global developmental delay, speech delay, febrile seizures, optic nerve hypoplasia, tube feeding, dysmorphic facial features | Limited Original Testing | ReNU syndrome (**#**620851) | Pathogenic | PS4, PM2, PP5 |
| PMGRC-151-151-0 | Syndromic | *GLMN* (#601749; NM_053274.3) | c.157_161del | p.(Glu52_Lys53insTer) | [NC_000001.11:g.92297411_92297415del](https://www.ncbi.nlm.nih.gov/nuccore/NC_000001.11?report=graph&search=NC_000001.11%3Ag.92297411_92297415del) | Unknown | Clinical diagnosis of glomuvenous malformations | Limited Original Testing | Glomuvenous malformations (**#**138000) | Pathogenic | PS4, PVS1, PM2 |
| PMGRC-158-158-0 | Syndromic | *COQ2* (#609825; NM_001358921.2) | c.421G>C,  c.138dup | p.(Val141Leu)  p.(Ala47fs) | [NC_000004.12:g.83273617C>G](https://www.ncbi.nlm.nih.gov/nuccore/NC_000004.12?report=graph&search=NC_000004.12%3Ag.83273617C%3EG)  [NC_000004.12:g.83284632dup](https://www.ncbi.nlm.nih.gov/nuccore/NC_000004.12?report=graph&search=NC_000004.12%3Ag.83284632dup) | Maternally inherited,  Paternally inherited | Retinal atrophy, horseshoe kidney, nephrotic syndrome | New Information/Reclassification | Coenzyme Q10 deficiency, primary, 1 (**#**607426) | LP  LP | PM3, PM2, PP3  PS4, PVS1, PM2, PM3 |
| PMGRC-170-170-0 | Neurodevelopmental | *GRIA4* (#138246; NM_000829.4) | c.1643G>C | p.(Trp548Ser) | NC_000011.10:g.105924565G>C | De novo | Hypotonia, feeding difficulties, global developmental delays of motor and language skills, microcephaly, delayed visual attention, abnormal brain MRI | New Information/Reclassification | Neurodevelopmental disorder with or without seizures and gait abnormalities (**#**617864) | LP | PM2, PP3, PS2, PP2 |
| PMGRC-175-175-0 | Syndromic | *PTPN11* (#176876; NM_002834.5) | c.1510A>G | p.(Met504Val) | [NC_000012.12:g.112489086A>G](https://www.ncbi.nlm.nih.gov/nuccore/NC_000012.12?report=graph&search=NC_000012.12%3Ag.112489086A%3EG) | De novo | Pulmonary valve stenosis, toe syndactyly, widely spaced nipples | Previously Known | Noonan syndrome 1 **(#**163950) | Pathogenic | PS3, PS4, PM6, PM1, PP2, PM2, PP3 |
| PMGRC-176-176-0 | Syndromic | *SCN8A* (#600702; NM_001330260.2) | c.2549G>A | p.(Arg850Gln) | [NC_000012.12:g.51765675G>A](https://www.ncbi.nlm.nih.gov/nuccore/NC_000012.12?report=graph&search=NC_000012.12%3Ag.51765675G%3EA) | Nonmaternal | Refractory seizures, epileptic encephalopathy, infantile spasms, global developmental delay, hypotonia, tube feeding, respiratory insufficiency, hypotonia | Previously Known | Developmental and epileptic encephalopathy 13 (**#**614558) | Pathogenic | PS2, PM1, PP2, PM2, PM5 |
| PMGRC-178-178-0 | Neurodevelopmental | *CHD1* (#602118; NM_001270.4) | c.1010C>T | p.(Thr337Ile) | NC_000005.10:g.98899555G>A | De novo | Motor delay, hypotonia, stereotypic behaviors, autism spectrum disorder | New Information/Reclassification | Pilarowski-Bjornsson syndrome (**#**617682) | LP | PM2, PS2, PP2 |
| PMGRC-204-204-0 | Neurodevelopmental | *FBXO31* (#609102; NM_024735.5) | c.1000G>A | p.(Asp334Asn) | [NC_000016.10:g.87334283C>T](https://www.ncbi.nlm.nih.gov/nuccore/NC_000016.10?report=graph&search=NC_000016.10%3Ag.87334283C%3ET) | Nonmaternal | Abnormal brain MRI, gross motor delay, speech delay, hypertonia, lower extremity spasticity, esotropia, mixed receptive-expressive language disorder | New Information/Reclassification | FBXO31-related spastic-dystonic cerebral palsy syndrome [28] | LP | PM3, PM2 |
| PMGRC-205-205-0 | Syndromic | *SLC6A8* (#300036; NM_005629.4) | c.1255-3_1255-2delCA | p.? | NC_000023.11:g.153694124CCA-C | Unknown | Acute liver failure, acute kidney injury, autism spectrum disorder | Limited Original Testing | Cerebral creatine deficiency syndrome 1 (**#**300352) | LP | PP4, PVS1, PM2 |
| PMGRC-212-212-0 | Other | *OCA2* (#611409; NM_000275.3) | c.1465A>G  a complex rearrangement with deep intronic breakpoints | p.(Asn489Asp)  p.? | [NC_000015.10:g.27983383T>C](https://www.ncbi.nlm.nih.gov/nuccore/NC_000015.10?report=graph&search=NC_000015.10%3Ag.27983383T%3EC)  hg19 NC_000015.9:g.[28119923_28303785del];[28337021_28339403delinsCCTGGTTGTAGGTCTAACCTGGTTAGAATCA;28143225_28285967inv;C] | Paternally inherited,  Maternally inherited | Oculocutaneous albinism | Limited Original Testing | Oculocutaneous albinism, type II (**#**203200) | Pathogenic  Pathogenic | PM2, PM1, PP3, PP5  PVS1, PP5 |
| PMGRC-220-220-0 | Syndromic | *HADHB* (#143450; NM_000183.3) | c.1390-515_1390-499del | p.? | NC_000002.12:g.26289403-26289469del | Maternally inherited | Hypoparathyroidism, LCHAD, mitochondrial trifunctional protein deficiency, pancytopenia, bone marrow deficiency, nephrotic syndrome, tube feeding | Limited Original Testing | Mitochondrial trifunctional protein deficiency 2 (#620300) [29] | LP | PM2, PM3, PVS1 |
| PMGRC-265-265-0 | Syndromic | *RNF220* (#616136; NM_018150.4) | c.1094G>A | p.(Arg365Gln) | [NC_000001.11:g.44636130G>A](https://www.ncbi.nlm.nih.gov/nuccore/NC_000001.11?report=graph&search=NC_000001.11%3Ag.44636130G%3EA) | Biparental homozygous | Bilateral sensorineural hearing loss, leukodystrophy, seizures, mixed receptive-expressive language disorder, photoreceptor degeneration, intention tremor | New Info/Reclassification | Leukodystrophy, hypomyelinating, 23, with ataxia, deafness, liver dysfunction, and dilated cardiomyopathy (**#**619688) | Pathogenic | PS4, PM2, PP3 |
| PMGRC-316-316-0 | Syndromic | *PTPN11* (#176876; NM_002834.5) | c.794G>A | p.(Arg265Gln) | [NC_000012.12:g.112472981G>A](https://www.ncbi.nlm.nih.gov/nuccore/NC_000012.12?report=graph&search=NC_000012.12%3Ag.112472981G%3EA) | Paternally inherited | Short stature, gross motor delays | Limited Original Testing | Noonan syndrome 1 **(#**163950) | Pathogenic | PS3, PS4, PP1, PS2, PM2, PP2, PM2, PM5 |
| PMGRC-332-332-0 | Neurodevelopmental | *CRPPA* (#614631; NM_001101426.4) | c.789+973C>G,  c.1120-1G>T | p.?  p.? | NC_000007.14:g.16307550G>C  NC_000007.14:g.16216198C>A | Maternally inherited,  Paternally inherited | Global developmental delay, hypotonia, chronic lung disease, retinopathy, severe optic atrophy, tube feeding, abnormal brain MRI | Limited Original Testing | Muscular dystrophy-dystroglycanopathy (congenital with brain and eye anomalies), type A, 7 (**#**614643) | LP  Pathogenic | PM3, PM2, PVS1  PM3, PS2, PVS1, PM2 |
| PMGRC-355-355-0 | Neurodevelopmental | *TCF7L2* (#602228; NM_001367943.1) | c.1156C>T | p.(Arg240Cys) | [NC_000010.11:g.113151879C>T](https://www.ncbi.nlm.nih.gov/nuccore/NC_000010.11?report=graph&search=NC_000010.11%3Ag.113151879C%3ET) | De novo | Walking difficulties, unsteady gait, lateral lisps, ADHD, distal weakness, hammertoes, gynecomastia, learning disabilities | New Information/Reclassification | TCF7L2- associated syndromic neurodevelopmental disorder [30] | LP | PM2, PP3, PM6 |
| PMGRC-366-366-0 | Neurodevelopmental | *DDX17* (#608469; NM_006386.5) | c.1077G>A | p.(Trp359*) | NC_000022.11:g.38494767C>T | De novo | Hypotonia, global developmental delay, gross motor delay, mixed receptive-expressive language disorder, foot drop, steppage gait, pes planus, ataxic gait | New Information/Reclassification | DDX17- associated neurodevelopmental disorder [31] | Pathogenic | PVS1, PM2, PS2 |
| PMGRC-388-388-0 | Syndromic | *RNU4-2* (#620823; NC_000012.12) | n.64_65insT | NA | [NC_000012.12:g.120291839_120291840insA](https://www.ncbi.nlm.nih.gov/nuccore/NC_000012.12?report=graph&search=NC_000012.12%3Ag.120291839_120291840insA) | De novo | Global developmental delay, hemiparesis, microcephaly, tube feeding, growth delays, truncal hypotonia, autism spectrum disorder, nonverbal, speech apraxia, sensory processing difficulties, abnormal brain MRI | Limited Original Testing | ReNU syndrome (**#**620851) | Pathogenic | PS4, PM2, PP5 |
| PMGRC-392-392-0 | Neurodevelopmental | *COQ4* (#612898; NM_016035.5) | c.718C>T,    c.202+4A>C | p.(Arg240Cys)  p. ? | [NC_000009.12:g.128333565C>T](https://www.ncbi.nlm.nih.gov/nuccore/NC_000009.12?report=graph&search=NC_000009.12%3Ag.128333565C%3ET" \t "_blank)  [NC_000009.12:g.128323151A>C](https://www.ncbi.nlm.nih.gov/nuccore/NC_000009.12?report=graph&search=NC_000009.12%3Ag.128323151A%3EC) | Maternally inherited,  Paternally inherited | Spastic diplegia, gross motor delays, speech articulation delay, unsteady gait, walking difficulties | New Information/Reclassification | Spastic ataxia 10, (**#**620666) | LP | PM3, PS3, PM2, PP3, PM5 |
| PMGRC-403-403-0 | Syndromic | *PEX1* (#602136; NM_000466.3) | c.1359+601A>G | p.? | NC_000007.14:g. 92513247T>C | Maternally inherited | Zellweger Spectrum Disorder, elevated very long-chain fatty acids, decreased plasmalogen | Limited Original Testing | Peroxisome biogenesis disorder 1A (**#**214100) | LP | PVS1, PM3, PM2, |
| PMGRC-418-418-0 | Neurodevelopmental | *ZIC2* (#603073; NM_007129.5) | c.1030_1032del | p.(Trp359*) | NC_000013.11:g.99983094_99983096del | De novo | Alobar holoprosencephaly | Limited Original Testing | Holoprosencephaly 5 (**#**609637) | LP | PM2, PM4, PM6 |
| PMGRC-437-437-0 | Syndromic | *GLI2* (#165230; NM_001374353.1) | c.1111_1120del | p.(Ile371fs) | NC_000002.12:g.120971992_120972001del | Paternally inherited | Septopreoptic holoprosencephaly, solitary median maxillary incisor syndrome, low thyroid, underdeveloped pituitary, sensory processing disorder, failure to thrive | Limited Original Testing | Holoprosencephaly 9 (**#**610829) | LP | PM2, PVS1 |
| PMGRC-440-440-0 | Syndromic | *ZIC2* (#603073; NM_007129.5) | c.1013del | p.(Pro338fs) | NC_000013.11:g.99983074TC>T | De novo | Deafness, cerebral palsy, holoprosencephaly | Previous testing information not provided | Holoprosencephaly 5 (**#**609637) | Pathogenic | PM2, PVS1, PS2 |
| PMGRC-451-451-0 | Syndromic | *ZIC2* (#603073; NM_007129.5) | c.1377_1406dup | p.(Ala461_Ala470dup) | [NC_000013.11:g.99985460_99985489dup](https://www.ncbi.nlm.nih.gov/nuccore/NC_000013.11?report=graph&search=NC_000013.11%3Ag.99985460_99985489dup) | De novo | Semilobar holoprosencephaly, diabetes insipidus, delayed development | Limited Original Testing | Holoprosencephaly 5 (**#**609637) | Pathogenic | PS4, PP1, PS3, PM2, PM4 |
| PMGRC-509-509-0 | Syndromic | *RNU4-2* (#620823; NC_000012.12) | n.64_65insT | NA | [NC_000012.12:g.120291839_120291840insA](https://www.ncbi.nlm.nih.gov/nuccore/NC_000012.12?report=graph&search=NC_000012.12%3Ag.120291839_120291840insA) | De novo | Microcephaly, hypotonia, delayed myelination, gross motor delay, brachycephaly, abnormal brain MRI, global developmental delay | Limited Original Testing | ReNU syndrome (**#**620851) | Pathogenic | PS4, PM2, PP5 |
| PMGRC-538-538-0 | Syndromic | *AGO2* (#606229; NM_012154.5) | c.602G>T | p.(Gly201Val) | [NC_000008.11:g.140560427C>A](https://www.ncbi.nlm.nih.gov/nuccore/NC_000008.11?report=graph&search=NC_000008.11%3Ag.140560427C%3EA) | Nonmaternal | Hip dysplasia, congenital hypotonia, cleft palate, global developmental delay, hearing loss, Pierre Robin sequence, seizures, speech delay, cerebral palsy, scoliosis, congenital heart defects, dysmorphic facial features | New Information/Reclassification | Lessel-Kreienkamp syndrome (**#**619149) | LP | PS4, PM2, PP3, PP2 |
| PMGRC-540-540-0 | Syndromic | *KMT2C* (#606833; NM_170606.3) | c.5668C>T | p.(Arg1890*) | NC_000007.14:g.152182192G>A | Maternally inherited | Hypotonia, fine motor delay, gross motor delay, sensory processing disorder, speech delay, decreased oral tone, selective mutism, ADHD, developmental coordination disorder, dysmorphic facial features | Limited Original Testing | Kleefstra syndrome 2 (**#**617768) | LP | PM2, PVS1 |
| PMGRC-569-569-0 | Neurodevelopmental | *STAG2* (#300826; NM_001042749.2) | 87kb deletion of chrX:123,946,386-124,003590  Exon 1-4 deletion | p.? | [NC_000023.11:g.124061012_124094911del](https://www.ncbi.nlm.nih.gov/nuccore/NC_000023.11?report=graph&search=NC_000023.11%3Ag.124061012_124094911del) | Unknown | Holoprosencephaly, global developmental delay, scoliosis, apraxia of speech, vision problems, vertebral anomaly | Previously Known | Holoprosencephaly-13, X-linked (#301043) | LP | 1A, 3A, 2E, 4L |
| PMGRC-599-599-0 | Neurodevelopmental | *RNU4-2* (#620823; NC_000012.12) | n.69C>T | NA | [NC_000012.12:g.120291835G>A](https://www.ncbi.nlm.nih.gov/nuccore/NC_000012.12?report=graph&search=NC_000012.12%3Ag.120291835G%3EA) | De novo | Small for gestational age, abnormal visual development/eye movement irregularity, global developmental delay, hypotonia, abnormal brain MRI | Limited Original Testing | ReNU syndrome (**#**620851) | LP | PS4, PM2, PS2 |
| PMGRC-635-635-0 | Syndromic | *CDK13* (#603309; NM_003718.5) | c.425dup | p.(Leu143fs) | NC_000007.14:g.39951066dup | Nonmaternal | Congenital heart disease, dysmorphic facial features, short stature, ADHD, global developmental delay | New Information/Reclassification | Congenital heart defects, dysmorphic facial features, and intellectual developmental disorder (**#**617360) | LP | PM2, PVS1 |
| PMGRC-658-658-0 | Syndromic | *DBT* (#248610; NM_001918.5) | c.1282-4218G>A | p.? | NC_000001.11:g. 100200640C>T | Biparental homozygous | Prenatal diagnosis of Argininosuccinic acid lyase deficiency and thiamine-responsive maple syrup urine disease | Limited Original Testing | Argininosuccinic aciduria, Maple syrup urine disease, type II (**#**620699) | LP | PM2, PVS1 |
| PMGRC-677-677-0 | Syndromic | *TPM3* (# 191030; NM_152263.4) | c.835C>G | p.(Leu279Val) | [NC_000001.11:g.154169324G>C](https://www.ncbi.nlm.nih.gov/nuccore/NC_000001.11?report=graph&search=NC_000001.11%3Ag.154169324G%3EC) | De novo | Pierre Robin sequence, tube feeding, gastroparesis, mannose-binding lectin deficiency, recurrent aspiration pneumonia and sinusitis, abnormal brain MRI, hypotonia | Limited Original Testing | Congenital Myopathy 4A, Autosomal Dominant (**#**255310) | LP | PP3, PM2, PP2, PM6 |
| PMGRC-700-700-0 | Syndromic | *OFD1* (#300170; NM_003611.3) | c.2413dup | p.(Gln805fs) | NC_000023.11:g.13762369dup | Maternally inherited | Semilobar holoprosencephaly, cleft lip and palate, bilateral postaxial polydactyly of hands and feet, bilateral cryptorchidism, Dandy-Walker malformation, ventriculomegaly, hypotelorism, seizures | Limited Original Testing | Joubert syndrome 10 (**#**300804) | LP | PVS1, PM2 |
| PMGRC-753-753-0 | Syndromic | *MSL2* (#614802; NM_018133.4) | c.535G>T | p.(Glu179*) | [NC_000003.12:g.136152346C>A](https://www.ncbi.nlm.nih.gov/nuccore/NC_000003.12?report=graph&search=NC_000003.12%3Ag.136152346C%3EA) | De novo | Gross motor delay, speech delay, hypotonia, microcephaly, frontal bossing, fine hair, ptosis, pectus excavatum, low muscle tone, Mongolian spot in lower back, global developmental delay | New Information/Reclassification | Karayol-Borroto-Haghshenas neurodevelopmental syndrome (**#**620985) | Pathogenic | PVS1, PM2, PP5 |
| PMGRC-812-812-0 | Syndromic | *NDST1* (#600853; NM_001543.5) | c.1850 C>T | p.(Thr617Ile) | NC_000005.10:g.150542851C>T | Biparental homozygous | Ankyloglossia, gross motor delays, thoracolumbar kyphosis, global developmental delay, axial hypotonia | New Information/Reclassification | Intellectual Developmental Disorder, Autosomal Recessive 46 (**#**616116) | VUS | PM2, PP3 |
| PMGRC-817-817-0 | Syndromic | *MT-TL1* (#590050; NC_012920.1) | n.14A>G | r.(?) | [NC_012920.1:m.3243A>G](https://www.ncbi.nlm.nih.gov/nuccore/NC_012920.1?report=graph&search=NC_012920.1%3Am.3243A%3EG) | Maternally inherited | Fatigue, headache, recurrent fever, tinnitus, recurrent episodes of infection, sudden loss of hearing | Limited Original Testing | Diabetes-deafness syndrome (**#**520000) | Pathogenic | PS4, PM2, PP5 |
| PMGRC-890-890-0 | Cardiovascular | *TTN* (#188840; NM_001267550.1) | c.59604_59607delAAAG | p.(Gly19871fs) | [NC_000002.12:g.178559330_178559333del](https://www.ncbi.nlm.nih.gov/nuccore/NC_000002.12?report=graph&search=NC_000002.12%3Ag.178559330_178559333del) | Unknown | Dilated Cardiomyopathy | Limited Original Testing | Dilated Cardiomyopathy, 1G (**#**604145) | Pathogenic | PS4, PVS1, PM2 |
| PMGRC-897-897-0 | Cardiovascular | *SCN5A* (#600163; NM_198056.3) | c.3911C>T | p.(Thr1304Met) | [NC_000003.12:g.38562467G>A](https://www.ncbi.nlm.nih.gov/nuccore/NC_000003.12?report=graph&search=NC_000003.12%3Ag.38562467G%3EA) | Unknown | Dilated Cardiomyopathy | Previously Known | Dilated Cardiomyopathy, 1E (**#**601154) | LP | PM2, PP3, PP5 |
| PMGRC-904-904-0 | Cardiovascular | *DMD* (#300377; NM_004006.3) | c.9851G>A | p.(Trp3284*) | [NC_000023.11:g.31182861C>T](https://www.ncbi.nlm.nih.gov/nuccore/NC_000023.11?report=graph&search=NC_000023.11%3Ag.31182861C%3ET) | Unknown | Dilated Cardiomyopathy | Previously Known | Dilated Cardiomyopathy, 3B (**#**302045) | Pathogenic | PS4, PVS1, PM2, PP5 |
| PMGRC-911-911-0 | Cardiovascular | *MYH6* (#160710; NM_002471.4) | c.2489C>T | p.(Pro830Leu) | [NC_000014.9:g.23394264G>A](https://www.ncbi.nlm.nih.gov/nuccore/NC_000014.9?report=graph&search=NC_000014.9%3Ag.23394264G%3EA) | Unknown | Dilated Cardiomyopathy | Limited Original Testing | Dilated Cardiomyopathy, 1EE (**#**613252) | Pathogenic | PP3, PM2, PP5 |
| PMGRC-914-914-0 | Cardiovascular | *TNNT2* (#191045; NM_001001430.3) | c.517C>T | p.(Arg173Trp) | [NC_000001.11:g.201363349G>A](https://www.ncbi.nlm.nih.gov/nuccore/NC_000001.11?report=graph&search=NC_000001.11%3Ag.201363349G%3EA) | Unknown | Dilated Cardiomyopathy | Limited Original Testing | Dilated Cardiomyopathy, 1D (**#**601494) | Pathogenic | PS4, PP1, PS3, PM1, PP2, PM2, PP5, PP3 |
| PMGRC-917-917-0 | Cardiovascular | *RBM20* (#613171; NM_001134363.3) | c.2497dupA | p.(Arg833fs) | NC_000010.11:g.110812894dup | Unknown | Dilated Cardiomyopathy | Limited Original Testing | Dilated Cardiomyopathy, 1DD (**#**613172) | Pathogenic | PM2, PVS1, PP4 |
| PMGRC-921-921-0 | Cardiovascular | *TNNI3K* (#613932; NM_015978.3) | c.2302G>A | p.(Glu768Lys) | [NC_000001.11:g.74492217G>A](https://www.ncbi.nlm.nih.gov/nuccore/NC_000001.11?report=graph&search=NC_000001.11%3Ag.74492217G%3EA) | Unknown | Dilated Cardiomyopathy | Limited Original Testing | Cardiac conduction disease with or without dilated cardiomyopathy (**#**616117) | LP | PS4, PP1, PS3, PM2 |
| PMGRC-991-991-0 | Connective tissue & Musculoskeletal disorders | *TGFBR1* (#190181; NM_004612.4) | c.1355del | p.(Pro452fs) | [NC_000009.12:g.99147753del](https://www.ncbi.nlm.nih.gov/nuccore/NC_000009.12?report=graph&search=NC_000009.12%3Ag.99147753del) | Unknown | Thoracic aortic aneurysm, Aortic dissection | Previous testing information not provided | Loeys-Dietz syndrome 1 (**#**609192) | LP | PM2, PVS1 |
| PMGRC-1040-1040-0 | Connective tissue & Musculoskeletal disorders | *FBN1* (#134797; NM_000138.5) | c.4096G>A | p.(Glu1366Lys) | [NC_000015.10:g.48474369C>T](https://www.ncbi.nlm.nih.gov/nuccore/NC_000015.10?report=graph&search=NC_000015.10%3Ag.48474369C%3ET) | Unknown | Thoracic aortic aneurysm, Aortic dissection | Previous testing information not provided | Marfan syndrome (**#**154700) | Pathogenic | PS2, PM1, PP2, PM2, PM5, PP3, PP5 |
| PMGRC-1091-1091-0 | Syndromic | *CREBBP* (#600140; NM_004380.2) | exon 2 deletion | p.? |  | De novo | Failure to thrive, global developmental delay, pectus excavatum, axial hypotonia, abnormal brain MRI, dysmorphic features | Limited Original Testing | Rubinstein-Taybi syndrome 1 (**#**180849) |  | 1A, 3A, 2E, 4L |

*ADHD* attention deficit hyperactivity disorder, *LCHAD* long-chain 3-hydroxyacyl-CoA dehydrogenase enzyme deficiency, *LP* likely pathogenic, *MRI* magnetic resonance imaging, *VUS* variant of uncertain significance

Supplemental Table 2. Candidate genes identified in unsolved cases. All genes are deposited in GeneMatcher. All variants in candidate genes are considered VUS until clinical validity is determined.

| **Participant ID** | **Candidate Gene (**MIM ID*, transcript ID) | **Variant (c.)** | **Variant (p.)** | **Variant (g.)** | **ACMG Classification** | **ACMG Criteria used** |
| --- | --- | --- | --- | --- | --- | --- |
| PMGRC-86-86-0 | *NRXN2* (#600566, NM_015080.4) | c.2107G>A | p.(Gly703Ser) | NC_000011.10:g.64660831C>T | VUS | PM2, PP3, PP2 |
| PMGRC-104-104-0 | *GALNT16* (#615132, NM_001168368.2) | c.502+1G>A | p.? | NC_000014.9:g.69325405 G>A | VUS | PM2, PP3, PS2 |
| PMGRC-116-116-0 | *ARHGAP21* (#609870, NM_020824.4)*; PDPK1* (#605213, NM_002613.5) | c.3692_3693del,  c.1588G>A | p.?  p.(Gly530Arg) | NC_000010.11:g.24595736_24595737del  NC_000016.10:g.2597684G>A | VUS  VUS | PM2, PVS1  PM2, PM6 |
| PMGRC-137-137-0 | *CSMD3* (#608399, NM_198123.2) | c.3158dup | p.(Arg1530*) | NC_000008.11:g.112650202dup | VUS | PM2, PVS1 |
| PMGRC-217-217-0 | *SLC16A13* ([HGNC:31037](https://www.genenames.org/data/gene-symbol-report/#!/hgnc_id/HGNC:31037), NM_201566.3)*; HIVEP1* (#194540, NM_002114.4)*; MYH7* (#160760, NM_000257.4) | c.410G>A,  c.395A>G,  c.6212dupA,  c.4187G>T | p.(Arg137Gln)  p.(Tyr132Cys)  p.(Tyr2071*)  p.(Arg1396Leu) | NC_000017.11:g.7038218G>A  NC_000017.11:g.7038203A>G  NC_000006.12:g.12130769dup  NC_000014.9:g.23417669C>A | VUS  VUS  VUS  VUS | PM2, PP3  PM2, PP3  PM2, PVS1  PM2, PP3, PM5, PP2 |
| PMGRC-239-239-0 | *FGD5* (#614788, NM_152536.4) | c.3372_3373del | p.(Gly1125Glufs*54) | NC_000003.12:g.14910896_14910897del | VUS | PM2, PVS1 |
| PMGRC-255-255-0 | *NRXN2* (#600566, NM_015080.4)*;*  *STRN3* (#614766, NM_001083893.2) | c.2605C>T;  c.1875C>A | p.(Arg869Trp)  p.(Tyr625*) | NC_000011.10:g.64651568G>A  NC_000014.9:g.30906890G>T | VUS  VUS | PM2, PP3, PP2  PM2 |
| PMGRC-279-279-0 | *VWA3B* (#614884, NM_144992.5) | c.59G>T,  c.1737+1G>A | p.(Gly20Val)  p.? | NC_000002.12:g.98093151G>T  NC_000002.12:g. 98194493G>A | VUS  VUS | PM2  PM2, PVS1 |
| PMGRC-291-291-0 | *NSL1* (#609174, NM_015471.4) | c.314-2_315delAGAT | p.? | NC_000001.11:g. 212784491AATCT>A | VUS | PM2, PM6 |
| PMGRC-312-312-0 | *TJP1* (#601009, NM_001330239.4) | c.3927-2_3927-1del | p.? | NC_000015.10:g. 29718067GT>G | VUS | PM2, PVS1 |
| PMGRC-363-363-0 | *CEP350* (#617870, NM_014810.5) | c.2258del | NC_000001.11:g.180020032del | p.(Gly753Glufs*11) | VUS | PM2, PVS1 |
| PMGRC-423-423-0 | *FAM193A* (#620037, NM_001366318.2)*;*  *MEX3D* (#611009)*;*  *CLPTM1* (#604783, NM_001294.4) | c.1390-2A>G;  c.595+1G>C;  c.953A>G | p.?  p.?  p.(Tyr318Cys) | NC_000004.12:g. 2659556A>G  NC_000019.10:g. 1567463C>G  NC_000019.10:g.44987338A>G | VUS  VUS  VUS | PM2, PVS1  PM2  PM2, PP3 |
| PMGRC-446-446-0 | *TP53BP2* (#602143, NM_001031685.3) | c.40G>A,  c.3240del | p.(Val14Met)  p.(Asp1080Glufs*5) | NC_000001.11:g.223821355C>T  NC_000001.11:g.223784238del | VUS  VUS | PM2  PM2 |
| PMGRC-485-485-0 | *HEATR1* (#620390, NM_018072.6)*;*  *PRR30* ([HGNC:28677](https://www.genenames.org/data/gene-symbol-report/#!/hgnc_id/HGNC:28677), NM_178553.4) | c.3613C>T,  c.275del,  c.-365-6C>T | p.(Gln1205*)  p.(Pro92Glnfs*33)  p. ? | NC_000001.11:g.236572505G>A  NC_000002.12:g.27138057del  NC_000002.12:g. 27138700G>A | VUS  VUS  VUS | PM2, PVS1  PM2  PM2, BP4 |
| PMGRC-525-525-0 | *PLXNC1* (#604259, NM_005761.3) | c.416del | p.(Leu139Argfs*4) | NC_000012.12:g.94149387del | VUS | PM2, PVS1 |
| PMGRC-746-746-0 | *DOP1A* (#616823, NM_015018.4)*; PCDH9* (#603581, NM_203487.3) | c.3559dup;  c.3291del | p.(Ile1187Asnfs*5)  p.(Ser1098Leufs*25) | NC_000006.12:g.83137601dup  NC_000013.11:g.66631260del | VUS  VUS | PM2  PM2 |
| PMGRC-772-772-0 | *MUC5B* (#600770, NM_002458.3) | c.15477+1G>C | p.? | NC_000011.10:g. 1254352G>C | VUS | PM2, PVS1 |
| PMGRC-782-782-0 | *CD177* (#162860, NM_020406.4) | c.99G>A | p.(Trp33*) | NC_000019.10:g.43353899G>A | VUS | PM2 |
| PMGRC-832-832-0 | *TLN2* (#607349, NM_015059.3)*; IL9* (#146931, NM_000590.2) | c.4645C>T;  c.183+6T>C,  c.202T>C | p.(Asp1549Tyr)  p.(Ser893Arg)  p.(Cys68Arg) | NC_000015.10:g.62761687G>T  NC_000005.10:g. 135895434A>G  NC_000005.10:g.135894133A>G | VUS  VUS  VUS | PM2, PM6, PP3  PM2, PP3  PM2 |
| PMGRC-949-949-0 | *GNRH2* (#602352, NM_178331.2) | c.213_214insTGTC | p.? | NC_000020.11:g.3044758_3044759insTGTC | VUS | PM2 |
| PMGRC-985-985-0 | *FMNL1* (#604656, NM_005892.4) | c.1379del | p.(Pro460Glnfs*19) | NC_000017.11:g.45241428del | VUS | PM2, PVS1 |
| PMGRC-992-992-0 | *PRR14L* (#621035, NM_173566.3) | c.4166del | p.(Tyr318Cys) | NC_000022.11:g.31713673del | VUS | PM2, PVS1 |
| PMGRC-999-999-0 | *SLC17A9* (#612107, NM_022082.4) | c.287C>G | p.(Tyr132Cys) | NC_000020.11:g.62957470C>G | VUS | PM2 |
| PMGRC-1017-1017-0 | *NXF3* (#300316, NM_022052.2) | c.1231G>T | p.(Glu411*) | NC_000023.11:g.103079463C>A | VUS | BS2, PVS1 |
| PMGRC-1030-1030-0 | *CORO1C* (#605269, NM_014325.4) | c.474G>A | p.(Trp158*) | NC_000012.12:g.108658894C>T | VUS | PM2, PVS1 |
| PMGRC-1048-1048-0 | *NEURL4* (#615865, NM_032442.3) | c.1606G>T | p.(Glu536*) | NC_000017.11:g.7325234C>A | VUS | PM2, PVS1 |
| PMGRC-1050-1050-0 | *CUL1* (#603134, NM_003592.3) | c.571_575del | p.(Arg191Trpfs*2) | NC_000007.14:g.148759584_148759588del | VUS | PM2, PVS1 |

*HGNC ID was used when MIM ID was not available, *LP* likely pathogenic, *VUS* variant of uncertain significance
